# Sensory Profile of Bipolar patients with a Neurodevelopmental Phenotype

**DOI:** 10.64898/2026.03.25.26349295

**Authors:** Elisabeth Palleau, Ilian Salmi, Kaina Ahamada, Mathieu Gilson, Catarina Silva, Hugo Pergeline, Raoul Belzeaux, Christine Deruelle, Antoine Lefrère

## Abstract

**Background:** Bipolar disorder (BD) is increasingly conceptualized as a heterogeneous condition with a neurodevelopmental phenotype (NDP) identifying a subgroup with early neurodevelopmental vulnerability and poorer clinical outcomes. Sensory processing (SP) abnormalities are a core feature of neurodevelopmental disorders but remain poorly characterized in BD and may reflect underlying neurodevelopmental liability. We examined whether NDP load is associated with specific SP alterations in euthymic BD patients and whether NDP-based stratification explains SP variability better than conventional BD subtype (BD 1/2).

**Methods:** We assessed 102 euthymic BD patients and 45 healthy controls (HC) using the Adolescent/Adult Sensory Profile (AASP). NDP load (0–3) was computed from nine clinical variables grouped into neonatal, comorbidity, and neurodevelopmental clusters; a median split defined BD without NDP (BD) and BD with NDP (BD-ND). Associations between NDP load and AASP quadrants were analyzed using Spearman correlations with FDR correction. Group differences (BD, BD-ND, HC) were assessed using Welch’s ANOVA and post-hoc tests. Nested and multivariable linear regressions examined whether NDP classification explained SP variance beyond BD subtype, adjusting for age, sex, anxiety, and residual mood symptoms.

**Results:** Higher NDP load correlated with greater low registration (ρ=0.35, *p*<0.001, *q*=0.004), sensory sensitivity (ρ=0.30, *p*=0.001, *q*=0.004), and sensation avoiding (ρ=0.23, *p*=0.014, *q*=0.040), but not sensation seeking. BD-ND showed higher low registration, sensory sensitivity, and sensation avoiding than BD and HC (all *qs*<0.01). NDP classification explained more SP variance than BD subtype; with robust associations after adjustment.

**Conclusions:** Sensory processing alterations in BD are dimensionally associated with neurodevelopmental load and more accurately captured by NDP-based stratification than diagnostic subtype. SP alterations may represent a transdiagnostic marker of neurodevelopmental liability within BD, supporting biologically informed stratification approaches.

## Introduction

Bipolar disorder (BD) is a complex and heterogeneous psychiatric condition characterized by alternating mood states, including mania, hypomania, and depression.

Although significant progress has been made, its pathophysiology remains poorly understood and the search for reliable biomarkers is still ongoing. This heterogeneity makes BD a challenging disorder to predict and manage. In an effort to refine the clinical understanding of BD, a neurodevelopmental phenotype (NDP) has recently been hypothesized (Lefrere et al., 2023, 2024). This approach highlights a specific subgroup of BD patients with a distinct neurodevelopmental vulnerability.

This data-driven classification approach identified nine clinical variables that were grouped into three clusters: neonatal, comorbidities, and neurodevelopmental, allowing for the calculation of an individual NDP load score. Moreover, patients with higher NDP load showed a worse prognosis and increased neurological soft signs. Notably, these individuals also exhibited a poorer response to lithium treatment (Lefrere et al., 2024). Nevertheless, the NDP construct still requires further validation and refinement within a stratification framework integrating dimensional neurodevelopmental markers. The rationale is that dimensional markers are continuous, multilevel measures such as graded sensoriality or neurodevelopmental phenotype severity, as compared to categorical DSM labels, that have to potential to better capture inter-individual heterogeneity and may improve prognostic and treatment-response stratification.

In this perspective, emerging evidence suggests that sensory processing (SP) abnormalities may represent a key feature of neurodevelopmental vulnerability. This hypothesis is grounded in prior studies suggesting that SP deficits are among the earliest neurodevelopmental disruptions and may thus represent early markers of atypical neurodevelopment (Germani et al., 2014). Sensory processing (SP), defined as the way individuals perceive, interpret, and respond to sensory stimuli, has gained growing interest in psychiatry due to its strong links with neurodevelopmental disorders (NDDs) such as autism spectrum disorder (ASD) and attention-deficit/hyperactivity disorder (ADHD). In these conditions, atypical SP is well-documented, contributing to emotional dysregulation and functional impairments (Dellapiazza et al., 2021; Robertson & Baron-Cohen, 2017). Furthermore, research indicates that SP abnormalities are not limited to classic NDDs but extend to other psychiatric disorders, including schizophrenia (Javitt & Freedman, 2015) and, more recently, BD (Engel-Yeger et al., 2016; van den Boogert et al., 2022). The relevance of SP in mental health research has been emphasized since the National Institute of Mental Health (NIMH) recently expanded its Research Domain Criteria (RDoC) framework by adding sensorimotor systems as a sixth domain (Harrison et al., 2019; Sanislow et al., 2019). A recent meta analysis explored SP across psychiatric disorders, supporting its role as a meaningful transdiagnostic dimension (van den Boogert et al., 2022).

However, despite this relevance, the study of SP in BD remains limited and inconsistent. Studies have reported altered SP patterns across mood states, including *sensory sensitivity*, *sensation avoidance*, and *low registration* (Engel-Yeger et al., 2018; Serafini et al., 2017). Yet, findings are contradictory, particularly in euthymic patients, and tend to focus on isolated sensory modalities (e.g., olfaction, visual perception) rather than adopting a comprehensive multidimensional approach (Lahera et al., 2016; Negoias et al., 2019; O’Bryan et al., 2014). Such inconsistencies likely reflect the marked heterogeneity of BD. Rather than constituting a uniform feature of the disorder, SP alterations may characterize specific subgroups. Given that atypical SP is a core feature of neurodevelopmental conditions, SP disruptions in BD may index an underlying neurodevelopmental liability. To date, no study has directly investigated the association between SP profiles and the NDP subtype, leaving a gap in understanding whether sensory alterations contribute to structuring BD heterogeneity.

Research on SP commonly uses the Adult/Adolescent Sensory Profile (AASP, Brown et al., 2001), based on Dunn’s Four-Quadrant Model (Dunn, 1997). This model classifies SP into four profiles, *low registration, sensation seeking, sensory sensitivity*, and *sensation avoiding*, based on how individuals respond to sensory input. Widely validated across neurodevelopmental and psychiatric populations, the model offers a useful framework for linking SP patterns to cognitive and emotional functioning (Brown et al., 2001; Dunn, 1997).

We hypothesize that the sensory profile of euthymic bipolar patients with higher NDP load, which defines this NDP, is distinct from those without such traits. The goal of the present study is thus to investigate whether bipolar patients with a higher NDP load exhibit distinct SP profiles compared to BD patients with a lower NDP load.. Establishing these differences can not only shed light on the neurobiological mechanisms underlying SP abnormalities, but more importantly, provide early detection strategies and more tailored therapeutic interventions for bipolar patients with the NDP. A secondary objective of this study is to assess whether this NDP-based stratification explains SP alterations more accurately than conventional diagnostic subtype (BD 1 and BD 2).

## Material and Methods

### Participants

The study included 102 patients (61 women, 41 men), aged 18 to 64 years old (*M*_age_ = 38,02, *SD* = 12,77), recruited via the FondaMental Academic Centers of Expertise for Bipolar Disorders (FACE-BD) of Marseille between May 2024 and March 2025, and 45 healthy control (HC) participants (27 women and 18 man), aged 18 to 64 years old (*M*_age_ = 18,82, *SD* = 13,34).

All patients had a clinical diagnosis of BD (type 1, 2 or Not Otherwise Specified (NOS)) based on the Diagnostic and Statistical Manual of Mental Diseases, Fifth (DSM-5; APA, 2013). All were received by a multidisciplinary team in the Expert Center where they completed an exhaustive routine screening procedure. Only euthymic patients, that is, those who have not experienced a mood episode in the past three months, were included (APA, 2013). In compliance with the International Society for Bipolar Disorders (ISBD), remission was defined as an absence or minimal symptoms of both depression and mania as measured by the Montgomery-Åsberg Depression Rating Scale (MADRS, *scores below 15*) and the Young Mania Rating Scale (YMRS, *scores below 12*, Montgomery & Åsberg, 1979; Young et al., 1978). Other exclusion criteria involved factors known to significantly impact cognitive functioning: history of neurological disease (e.g., epilepsy or stroke), hearing and visual disabilities, electroconvulsive therapy in the past 12 months. Additional clinical information included the BD type and anxiety, measured by the State-Trait Anxiety Inventory (STAI, (Spielberger, C. D., et al., 1983).

The healthy control (HC) participants were recruited during the same period, from May 2024 to March 2025, from non-psychiatric healthcare and the administrative staff via posters displayed in strategic areas (e.g., break rooms, information boards) at Sainte-Marguerite Hospital in Marseille. Inclusion criteria included absence of known personal or family history of psychiatric or neurological disorders.

All participants provided their informed consent. This study was approved by the Ethics Committee of Aix-Marseille University (ref. no. 2025-03-13-09). The FondaMental Advanced Centers of Expertise for Bipolar Disorder (FACE-BD) cohort, as described by Lefrère et al. (2024) in their study defining the neurodevelopmental sub-phenotype of BD, was approved by the CPP-Ile de France IX on January 18, 2010. Sample sizes and data completeness for all clinical and sensory variables are detailed in Supplementary Table S1.

### Procedures

All patients first completed the standard clinical screening procedures from the Expert Center, and the NDP load was calculated for all of them (Lefrère et al., 2024). The HC group completed a brief sociodemographic and a clinical interview. All participants then completed the Adult/Adolescent Sensory Profile (AASP, Brown et al., 2001).

## Materials

### Neurodevelopmental phenotype (NDP) assessment

The NDP load was assessed using a screening tool developed by Lefrère et al. (2024), designed to capture cumulative markers of neurodevelopmental vulnerability. This load is based on nine clinical variables divided into three clusters : (1) *the neonatal cluster* (maternal age of 35 years or older; paternal age of 40 years or older); (2) *the comorbidities cluster* (onset of substance use, eating and anxiety disorders before the age of 16, onset on BD before the age of 18); and (3) *the neurodevelopmental cluster* (severe childhood trauma, as measured by the Childhood Trauma Questionnaire (CTQ, Bernstein et al., 2011), ADHD (clinical diagnosis or determined by scores over 46 at the Wender Utah Rating Scale (WURS, Ward, M. F, 1993), or specific learning disorders (e.g., dyslexia, dyspraxia). Each variable is scored 1 if present and 0 if absent, with a total score ranging from 0 to 3, reflecting the cumulative neurodevelopmental load. Higher scores indicate a higher neurodevelopmental load. For categorical analyses, BD patients were classified into two subgroups using a median split of the NDP load: bipolar disorder without neurodevelopmental phenotype (BD) and bipolar disorder with the neurodevelopmental phenotype (BD-ND) (Supplemental material, Figure 1).

**Figure 1.**
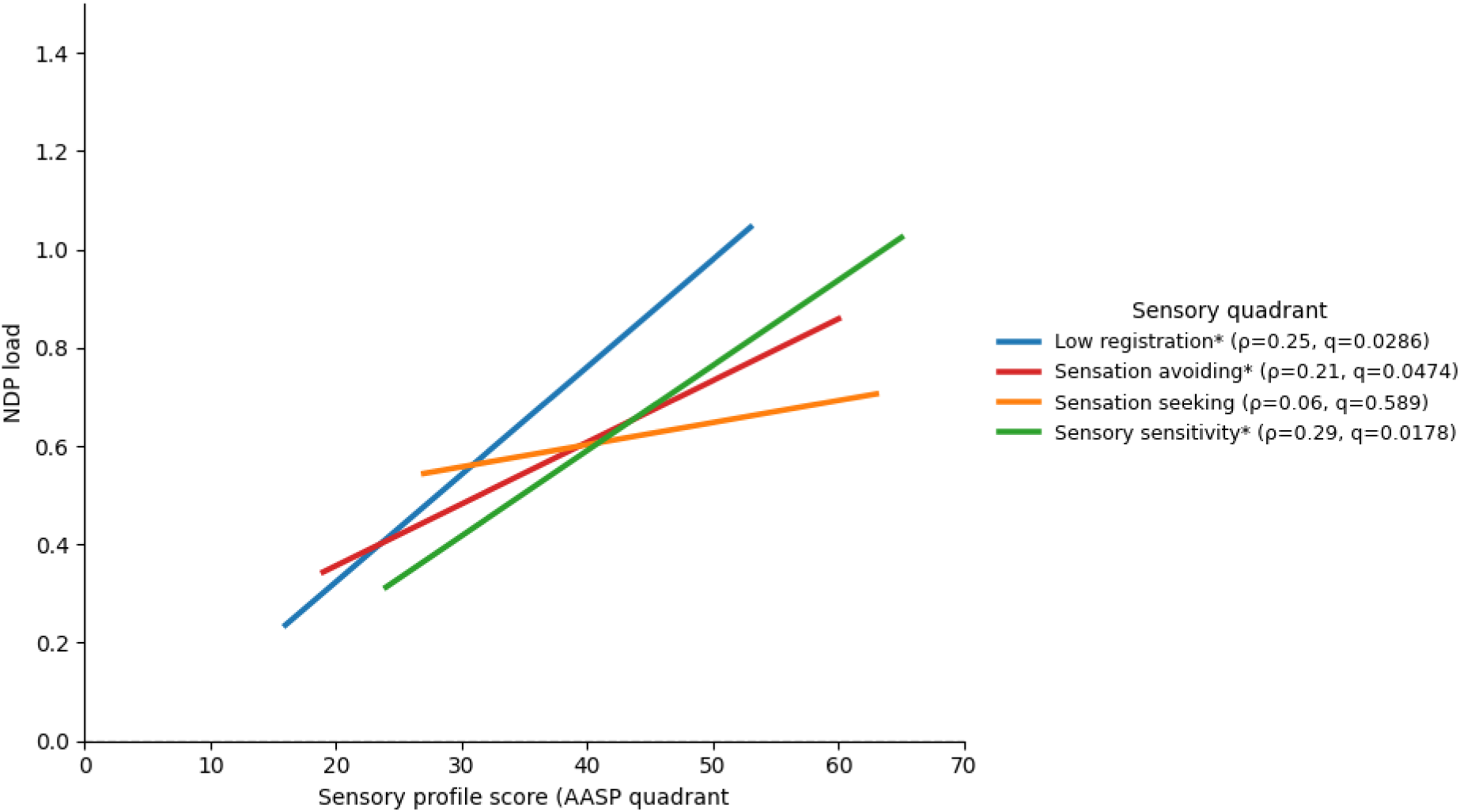
Associations between sensory processing quadrants and the NDP load in bipolar disorder. Associations between the Adolescent/Adult Sensory Profile (AASP) quadrant scores and the NDP load are shown in participants with bipolar disorder (BD and BD-ND). Lines represent least-squares fits for visualization purposes. Correlations were assessed using Spearman’s rank coefficients (ρ), with false discovery rate (FDR) correction applied across the four quadrants. Significant associations after FDR correction are indicated by an asterisk (*). Regression lines are displayed over the observed range of each quadrant score.

### Sensory profile assessment

Sensory profiles were assessed using the AASP (Brown et al., 2001). The AASP is a 60-item self-report questionnaire widely used to evaluate SP patterns. Participants indicate the frequency of their behavior responses to sensory experiences in daily-life on a 5-point Likert-type scale (ranging “*from almost never*” to “*almost always*”). Responses are organized based on Dunn’s theoretical model (Dunn, 1997), according to which four sensory quadrants are hypothesized: (1) *low registration* (difficulty detecting sensory input), (2) *sensation seeking* (active pursuit of sensory stimulation), (3) *sensory sensitivity* (heightened and passive reactions to sensory stimuli), and (4) *sensation avoiding* (active efforts to reduce sensory exposure). Standardized scores are then used to classify individuals in one out five sensory patterns, ranging from “*much less than most people*” to “*much more than most people*,” indicating the frequency of sensory behaviors relative to the general population. The Figure 2 (See Supplemental material) provides a visual summary of Dunn’s SP model.

**Figure 2.**
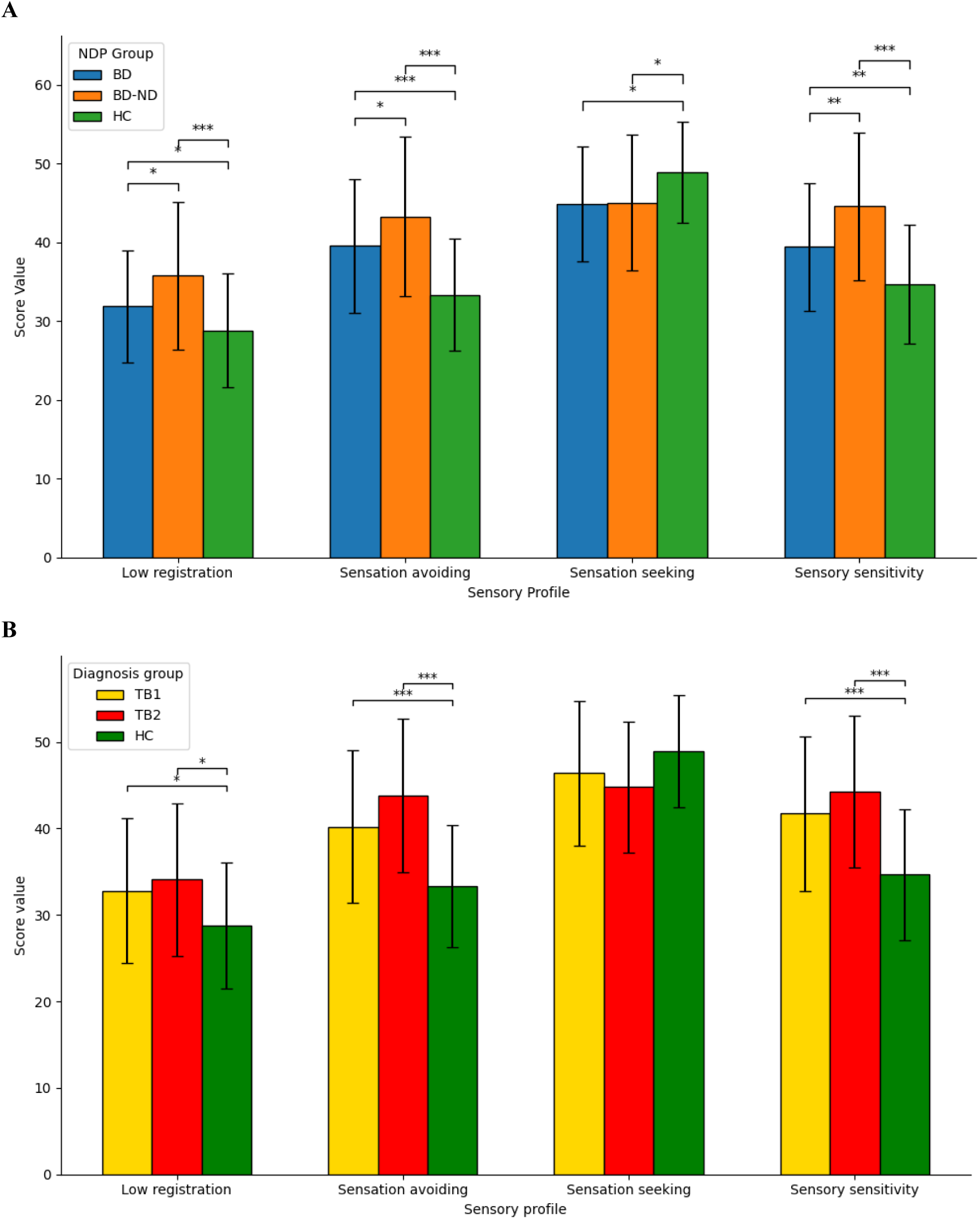
Sensory processing profiles across NDP and diagnostic groups. Mean (± SD) scores on the four AASP quadrants (Low registration, Sensation avoiding, Sensation seeking, Sensory sensitivity) are shown for BD, BD-ND, and HC (top panel), and for BD1, BD2, and HC (bottom panel). Asterisks indicate significant post-hoc comparisons (* q < 0.05, ** q < 0.01, *** q < 0.001) corresponding to pairwise Welch t-tests with false discovery rate (FDR) correction; the 3-group differences were tested using Welch ANOVA and are reported in the main text. Error bars represent standard deviations across the subjects.

### Statistical analysis

Statistical analyses were performed using Python (version 3.13) with libraries scipy.stats (version 1.16.3) and statsmodels (version 0.14.6). Descriptive statistics were used to summarize demographic, clinical, and sensory characteristics.

Associations between AASP quadrant scores and the continuous NDP load score were examined in bipolar participants using Spearman rank correlations. False discovery rate (FDR) correction (Benjamini–Hochberg) was applied across the four sensory quadrants. Categorical group comparisons were conducted across BD without predominant neurodevelopmental features (BD group), BD with neurodevelopmental features (BD-ND group), and healthy controls (HC group). Continuous variables were analyzed using Welch’s ANOVA for three-group comparisons and Welch’s t-tests for pairwise comparisons when HC data were unavailable (i.e. BD vs BD-ND). No imputation procedure was performedCategorical variables were compared using χ² tests. Post-hoc pairwise comparisons were performed using Welch t-tests with Benjamini–Hochberg FDR correction, as reported by the corrected significance value q in the text. To determine whether NDP-based classification provided explanatory value beyond conventional diagnostic subtype (BD1 vs BD2 types), we estimated a set of nested linear regression models separately for each AASP sensory quadrant (Low Registration, Sensation Seeking, Sensory Sensitivity, Sensation Avoiding). In all models, the sensory quadrant score was treated as a continuous outcome. First, a diagnosis-only model was fitted: Y ∼ Diagnosis (coded as TB1 vs TB2). Second, an NDP-only model was fitted: Y ∼ NDP group (BD vs BD-ND). Third, an additive model was fitted: Y ∼ Diagnosis + NDP group. Finally, an interaction model was fitted: Y ∼ Diagnosis × NDP group. Model fit was summarized using the coefficient of determination (R²) for each model. The incremental contribution of NDP beyond diagnosis was quantified as ΔR² between the additive model and the diagnosis-only model. The added value of the interaction term was quantified as ΔR² between the interaction model and the additive model. Nested model comparisons were evaluated using F-tests for changes in explained variance. In addition, model parsimony was examined using information criteria (AIC/BIC) as part of sensitivity analyses. Because heteroskedasticity was expected in clinical data, regression estimates were computed with heteroskedasticity-consistent (HC3) robust standard errors (as implemented in statsmodels library for linear regression models, “ols”). Finally, **as part of sensitivity analyses,** multivariable linear regression models were conducted in bipolar participants to assess the robustness of associations between the NDP load and sensory quadrants after taking confound factors into account, namely age, anxiety (STAI), depressive symptoms (MADRS), manic symptoms (YMRS), and sex. Outcomes were standardized, and heteroskedasticity-consistent (HC3) robust standard errors were applied. All analyses were conducted using complete-case approaches specific to each model. For each analysis, only participants with non-missing data on the variables included in the corresponding model (outcome and predictors) were retained. Statistical significance was set at p < .05 (two-tailed).

## Results

The demographic and clinical characteristics of the BD sample are summarized in Table 1. Participants were euthymic at assessment, as reflected by low mean scores on the MADRS (4.9 ± 4.2) and YMRS (1.6 ± 3.1). Group-level demographic, clinical, and sensory characteristics across the BD group, the BD-ND group, and the HC group are presented in Table 2.

**Table 1.** Clinical and sensory characteristics of the bipolar disorder sample.

| Variable | N | Mean (SD) |
| --- | --- | --- |
| Age (years) | 102 | 38.0 (12.8) |
| NDP load | 102 | 0.64 (0.53) |
| MADRS score | 102 | 4.9 (4.2) |
| YMRS score | 94 | 1.6 (3.1) |
| STAI score | 102 | 35.7 (11.7) |
| Low registration | 98 | 33.8 (8.5) |
| Sensation seeking | 98 | 45.0 (7.9) |
| Sensory sensitivity | 98 | 42.0 (9.1) |
| Sensation avoiding | 98 | 41.4 (9.5) |
| **Sex** |  |  |
| Female | 61 (59.8%) |  |
| Male | 41 (40.2%) |  |
| **Diagnosis** |  |  |
| TB1 | 37 (49.3%) |  |
| TB2 | 32 (42.7%) |  |
| TBNS | 5 (6.7%) |  |
| TDM | 1 (1.3%) |  |
| Missing | 27 (26.5%) |  |

Continuous variables are presented as mean (standard deviation) and categorical variables as n (%). N refers to the number of participants with available data for each variable.
*Age* is reported in years. *NDP load* reflects the dimensional neurodevelopmental polarity score. MADRS = Montgomery–Åsberg Depression Rating Scale; YMRS = Young Mania Rating Scale; STAI = State–Trait Anxiety Inventory. Sensory profiles (Low registration, Sensation seeking, Sensory sensitivity, Sensation avoiding) were assessed using the Adolescent/Adult Sensory Profile. Missing indicates participants without available diagnosis data.

**Table 2.**
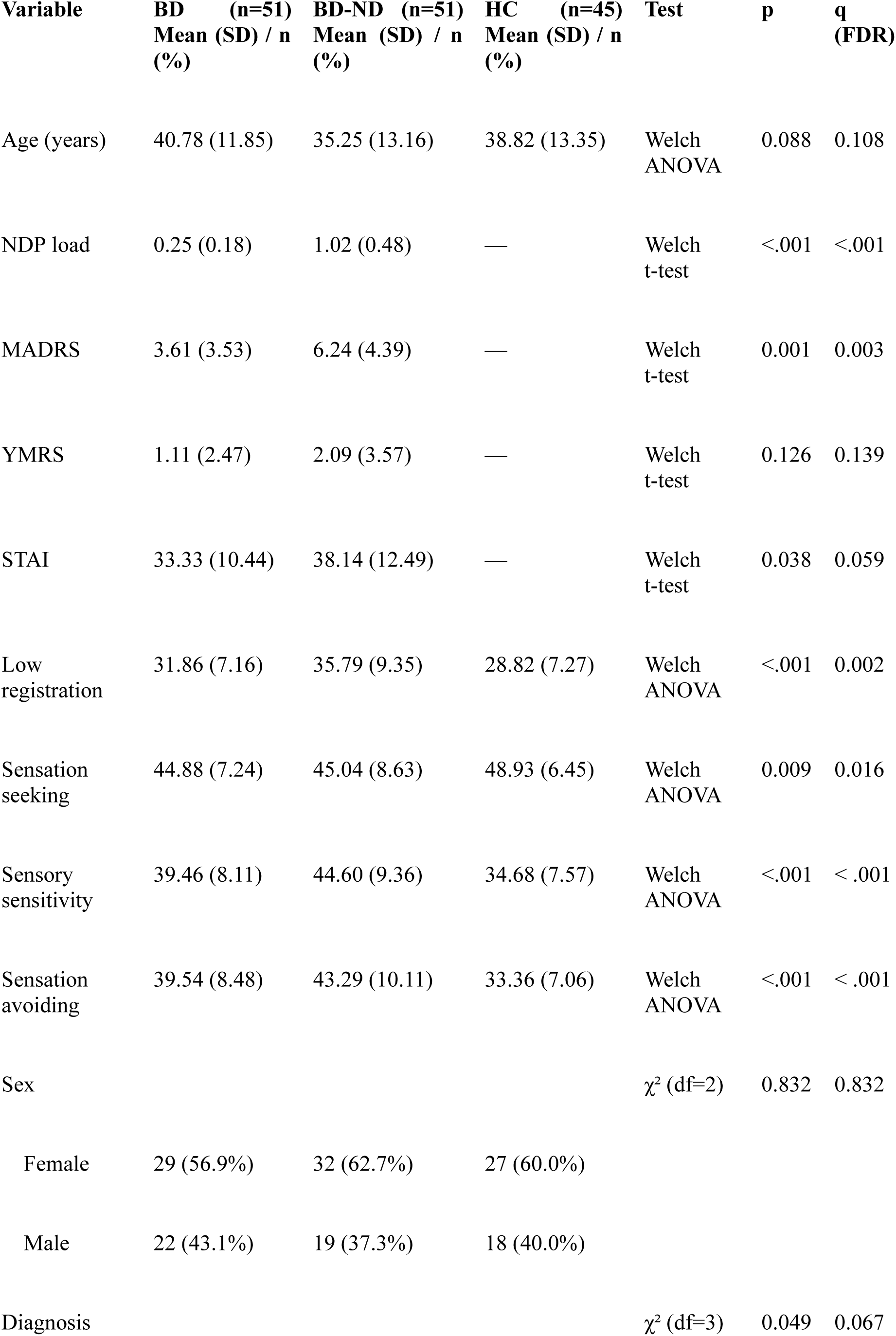
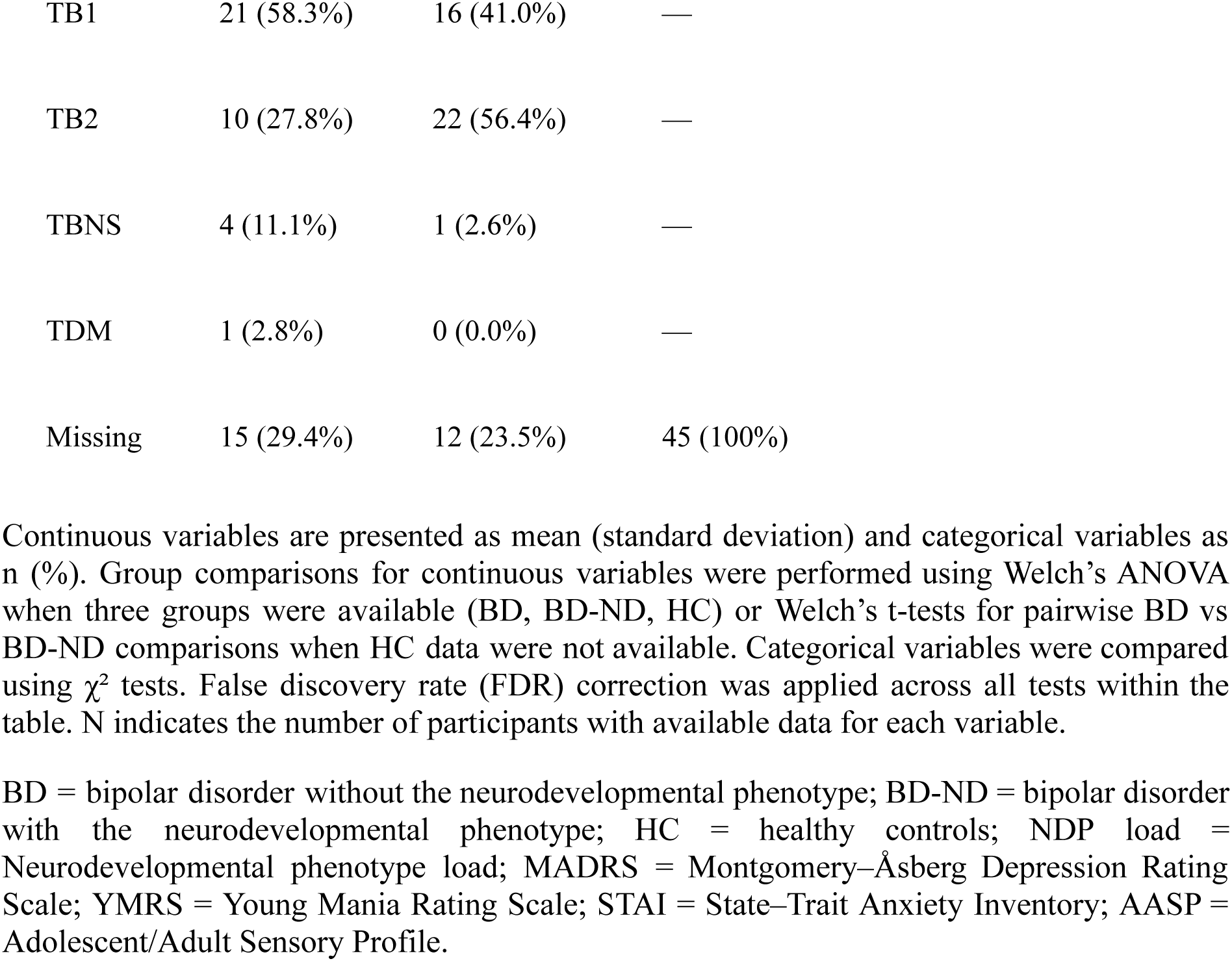
Clinical, sensory, and demographic characteristics across BD, BD-ND, and healthy controls.

| Variable | BD (n=51)<br>Mean (SD) / n<br>(%) | BD-ND (n=51)<br>Mean (SD) / n<br>(%) | HC (n=45)<br>Mean (SD) / n<br>(%) | Test | p | q<br>(FDR) |
| --- | --- | --- | --- | --- | --- | --- |
| Age (years) | 40.78 (11.85) | 35.25 (13.16) | 38.82 (13.35) | Welch<br>ANOVA | 0.088 | 0.108 |
| NDP load | 0.25 (0.18) | 1.02 (0.48) | — | Welch<br>t-test | <.001 | <.001 |
| MADRS | 3.61 (3.53) | 6.24 (4.39) | — | Welch<br>t-test | 0.001 | 0.003 |
| YMRS | 1.11 (2.47) | 2.09 (3.57) | — | Welch<br>t-test | 0.126 | 0.139 |
| STAI | 33.33 (10.44) | 38.14 (12.49) | — | Welch<br>t-test | 0.038 | 0.059 |
| Low<br>registration | 31.86 (7.16) | 35.79 (9.35) | 28.82 (7.27) | Welch<br>ANOVA | <.001 | 0.002 |
| Sensation<br>seeking | 44.88 (7.24) | 45.04 (8.63) | 48.93 (6.45) | Welch<br>ANOVA | 0.009 | 0.016 |
| Sensory<br>sensitivity | 39.46 (8.11) | 44.60 (9.36) | 34.68 (7.57) | Welch<br>ANOVA | <.001 | <.001 |
| Sensation<br>avoiding | 39.54 (8.48) | 43.29 (10.11) | 33.36 (7.06) | Welch<br>ANOVA | <.001 | <.001 |
| Sex | | | | $\chi^2$ (df=2) | 0.832 | 0.832 |
| Female | 29 (56.9%) | 32 (62.7%) | 27 (60.0%) |  |  |  |
| Male | 22 (43.1%) | 19 (37.3%) | 18 (40.0%) |  |  |  |
| Diagnosis | | | | $\chi^2$ (df=3) | 0.049 | 0.067 |

|  |  |  |  |
| --- | --- | --- | --- |
| TB1 | 21 (58.3%) | 16 (41.0%) | — |
| TB2 | 10 (27.8%) | 22 (56.4%) | — |
| TBNS | 4 (11.1%) | 1 (2.6%) | — |
| TDM | 1 (2.8%) | 0 (0.0%) | — |
| Missing | 15 (29.4%) | 12 (23.5%) | 45 (100%) |

Continuous variables are presented as mean (standard deviation) and categorical variables as n (%). Group comparisons for continuous variables were performed using Welch's ANOVA when three groups were available (BD, BD-ND, HC) or Welch's t-tests for pairwise BD vs BD-ND comparisons when HC data were not available. Categorical variables were compared using $\chi^2$ tests. False discovery rate (FDR) correction was applied across all tests within the table. N indicates the number of participants with available data for each variable.
BD = bipolar disorder without the neurodevelopmental phenotype; BD-ND = bipolar disorder with the neurodevelopmental phenotype; HC = healthy controls; NDP load = Neurodevelopmental phenotype load; MADRS = Montgomery-Åsberg Depression Rating Scale; YMRS = Young Mania Rating Scale; STAI = State-Trait Anxiety Inventory; AASP = Adolescent/Adult Sensory Profile.

### Associations between continuous NDP load and AASP quadrants

First, we analysed the NDP load as a continuous score and SP dimensions in the bipolar sample (Figure 1; Supplementary Figure S3). Higher NDP load was significantly associated with increased *low registration* (ρ = 0.35, p < 0,001, q = 0.004), *sensory sensitivity* (ρ = 0.30, p = 0,001, q = 0.004), and *sensation avoiding* (ρ = 0.23, p = 0,014, q = 0.040). In contrast, *sensation seeking* was not associated with NDP load (ρ = 0.07, p = 0,46, q = 0.470). These findings indicate that increased neurodevelopmental load is significantly associated with heightened sensory hypo-responsivity and sensory over-responsivity, but not to sensory seeking behaviors.

### Group comparisons (BD, BD-ND, HC, BD1 and BD2)

Secondly, we conducted a categorical analysis, exploring group differences in SP patterns. Based on their NDP load (median split method), two BD subgroups were determined: the *BD-ND group*, with a NDP, characterized by a high NDP load (NDP > 0.5), (32 women, 19 men, *M*_age_ = 35.25, *SD* = 13.16, ranging from 18 to 64 years old), and the *BD group*, without those features (29 women, 22 men, *M*_age_ = 40.78, *SD* = 11.85, ranging from 18 to 63 years old), see Table 2; Figure 2A. This analysis revealed significant group effects for *low registration* (p < .001), *sensory sensitivity* (p < .001), *sensation avoiding* (p < .001), but not regarding *sensation seeking* (p = .009). Post-hoc comparisons (Supplementary Table S2A) demonstrated that the BD-ND group exhibited higher *low registration, sensory sensitivity,* and *sensation avoiding* compared to both the BD group and the HC group (all qs < .01). *Sensation seeking* was significantly elevated in both BD groups compared to the HC group (q < 0,5), and no significant difference was observed between the BD and BD-ND groups. Stratification by diagnostic subtype (BD1 vs BD2), produced no differences across all four sensory quadrants (Figure 2B; Supplementary Table S2B). In contrast, both BD groups differed significantly from HC for *low registration, sensory sensitivity, and sensation avoiding* (all qs < .05).

### Comparative explanatory value of NDP and diagnostic models

The previous analyses showed that the NDP load had the potential to predict partly the SP measurements in three of the four quadrants, although causality from NDP to SP would require a deeper analysis with the temporal unfolding of symptoms, which is beyond our scope here. To further examine whether NDP-based classification provides explanatory value beyond conventional diagnostic subtype, we estimated nested linear regression models separately for each sensory quadrant (Figure 3). Overall, both diagnosis-only and NDP-only models explained relatively small proportions of variance in sensory processing (R² values ranging from 0.006 to 0.045). For Low Registration and Sensory Sensitivity, NDP-only models showed greater explanatory power than diagnosis-only models (Low Registration: R² = 0.045 vs 0.006; Sensory Sensitivity: R² = 0.037 vs 0.021). In contrast, for Sensation Avoiding, diagnostic subtype accounted for more variance than NDP classification (R² = 0.041 vs 0.008). Sensation Seeking was minimally explained by either framework (R² ≤ 0.013). When both predictors were entered simultaneously, the increase in explained variance remained modest (ΔR² ≤ 0.040 across quadrants), and adding an interaction term (Diagnosis × NDP) did not meaningfully improve model fit (ΔR² ≤ 0.007). Taken together, these findings suggest that neurodevelopmental polarity captures a meaningful but quantitatively modest portion of variability in sensory processing. Its explanatory contribution is comparable to, and in some domains exceeds, that of traditional bipolar diagnostic subtype. However, combining both classification approaches provides limited additional gain, indicating that their effects are largely overlapping rather than synergistic.

**Figure 3.**
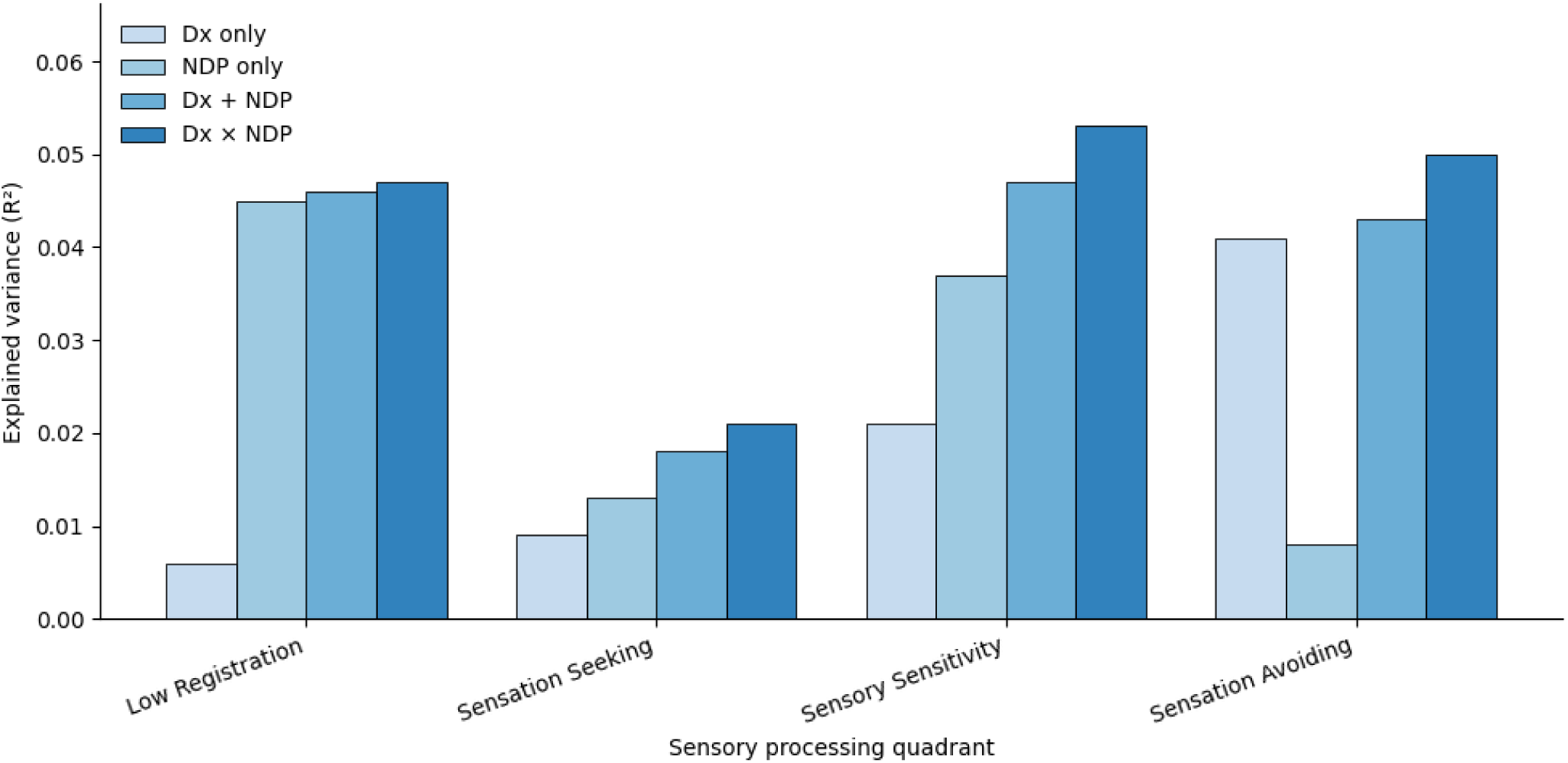
Explained variance of sensory processing profiles by diagnostic and NDP-based models. For each AASP quadrant (Low registration, Sensation seeking, Sensory sensitivity, Sensation avoiding), the proportion of explained variance (R²) is shown for four regression models: diagnosis only (Dx), NDP classification only (NDP), additive model (Dx + NDP), and interaction model (Dx × NDP). All models were estimated in the bipolar disorder sample using standardized outcomes. Higher R² values indicate greater explanatory power of the corresponding classification framework., For each sensory quadrant, the model with the lowest Bayesian Information Criterion (BIC) is indicated with an arrow, reflecting the most parsimonious model. Values above bars represent model R² estimates.

### Sensitivity analysis

Because the impact of mood and anxiety states on sensoriality is well established, we conducted a sensitivity analysis to ensure the robustness of our findings, accounting for potential residual affective and anxiety symptoms despite recruitment during euthymic states, in the absence of clinically characterized depressive or hypomanic/manic episodes. Multivariable regression analyses adjusting for age, anxiety (STAI), depressive symptoms (MADRS), and manic symptoms (YMRS) are presented in Supplementary Table S1. The associations between the NDP load and *low registration* (*β* = 0,25, 95% CI [0,06; 0,44], *p* = .01), *sensory sensitivity* (β = 0,23, 95% CI [0.02; 0,43], *p* = .03) remained statistically significant. The association between the NDP load and *sensation avoiding* became non-significant after adjustment (β = 0,18, 95% CI [-0.03; 0.39], *p* = .09). No association was found to be significant between the NDP load and *sensation seeking* after adjustment (*β* = 0,15, 95% CI [-0.07; 0.36], *p* = .2).

Taken together, both continuous and categorical analyses indicate that the NDP is robustly associated with specific SP alterations in BD, characterized by increased low registration and sensory sensitivity (with weaker evidence for sensation avoidance). Furthermore, the NDP-based explains SP alterations better than conventional diagnostic subtype.

## Discussion

This study investigated sensory processing in euthymic individuals with bipolar disorder and evaluated the efficiency of NDP as a stratification framework within BD. To our knowledge, this study represents one of the largest investigations to date of sensory processing alterations in euthymic individuals with BD.

The findings indicate that higher expression of the NDP, operationalized as NDP load, is significantly associated with increased *low registration*, *sensory sensitivity* and *sensation avoiding* behaviors. These results are consistent with previous research demonstrating similar sensory processing patterns in other neurodevelopmental and psychiatric disorders (e.g., van den Boogert et al., 2022). In particular, *low registration* and *sensory sensitivity* remained significantly associated with the NDP even after adjusting for anxiety and residual mood symptoms, which are well-known contributors to altered SP (Engel-Yeger & Dunn, 2011). Given that these associations were observed in euthymic patients, our results support the hypothesis that this specific sensory processing profile may constitute stable, trait-like features associated with the NDP in BD. Moreover, the observed gradient in sensory alterations provides empirical support for conceptualizing BD along a dimensional NDP continuum, with sensory processing contributing to clinically meaningful stratification within the disorder.

Complementing our dimensional analyses, categorical analyses showed that three sensory quadrants (*low registration, sensory sensitivity and sensation avoiding*) were elevated in BD-ND groups relative to HC participants. Conversely, both BD and BD-ND patients demonstrated a significant reduction in *sensation seeking* compared to HC participants. This aligns with Engel-Yeger and colleagues (2016) who reported reduced *sensation seeking* in BD.

Another key contribution of this study is empirical support for the NDP as a clinically meaningful stratifying framework in BD. Specifically, we demonstrated that NDP-based stratification captures variance in sensory phenotypes that conventional diagnostic subtype (BD 1 vs BD 2) does not. Across nested regression models and variance-explained comparisons, NDP classification accounted for more of the between-subject variability in sensory processing quadrants than did conventional diagnostic subtype (BD 1 vs BD 2), and additive or interaction models offered only marginal improvements. These findings suggest that the NDP indexes developmental liabilities, including early adversities, ADHD, learning disorders, and related markers (Lefrère et al., 2024), that are more directly linked to sensory integration and modulation processes than episode-based diagnostic labels. In practical terms, NDP-based stratification may identify a subgroup of patients who share convergent developmental risk and sensory dysregulation, thereby improving phenotypic homogeneity for biological and neuroimaging investigations.

This stratification framework has several neurobiological and clinical implications. From a neurobiological perspective, the co-occurrence of low registration and sensory sensitivity/avoidance may reflect disrupted gating and modulation: some individuals under-detect weak stimuli but over-react to salient inputs, consistent with models of atypical sensory integration (Miller et al., 2007) and with neuroimaging evidence implicating white-matter and thalamocortical circuitry in sensory dysfunction (Owen et al., 2013). From a clinical perspective, NDP screening could facilitate more precise stratification. Identifying patients with convergent developmental risk and sensory dysregulation may guide targeted assessments (e.g., occupational therapy, sensory modulation interventions) and enable neurobiological studies (neuroimaging, genomics), thereby enhancing power to detect biological correlates and treatment-response predictors (Lefrère et al., 2024).

The reduced sensation seeking sensory profile across BD calls for further comment. While some studies have reported elevated sensation seeking in manic or symptomatic samples (Aghaeimazraji et al., 2024; Parker, 2014), findings from our euthymic cohort together with prior meta-analytic work suggest that everyday sensory seeking may be attenuated as a trait in BD (van den Boogert et al., 2022). Such attenuation may contribute to reduced social engagement and adaptive functioning (Engel-Yeger et al., 2016; Escelsior et al., 2023). This pattern may carry meaningful clinical implications, mostly in relation to emotional regulation and social engagement. Previous studies suggest that *sensation seeking* may have a protective role (Engel-Yeger et al., 2016; Escelsior et al., 2023), possibly contributing to more adaptive emotional regulation of sensory input. Increased *sensation seeking* has been associated with greater social engagement, higher community participation and recovery in adults with severe mental illness (Forsberg et al., 2025; Pfeiffer et al., 2014), helping to reduce isolation among older persons (Engel-Yeger & Rosenblum, 2017). This implies that its diminution in BD could contribute to chronic functional impairments and should be considered in rehabilitation planning. Escelsior et al., 2023, found increased white matter integrity in the same structures associated with visuospatial processing in healthy adults with a sensation seeking profile. These results underscore the value of neuroimaging tools such as diffusion tensor imaging (DTI) in delineating sensory sensory phenotypes and distinguishing SP related alterations from overlapping conditions like autism and ADHD. These approaches may also prove informative in characterizing sensory traits within the NDP dimension in BD.

Clinically, integrating NDP screening and SP assessment into routine evaluation may have several applications. First, atypical sensory responses often appear early in development, with some signs observable as early as six months of age in children later diagnosed with ASD (Germani et al., 2014). They may serve as premorbid indicators of psychiatric vulnerability, especially in individuals with a pronounced NDP profile. Second, including sensory assessments in clinical screening may help identify the NDP subgroup of patients earlier and support more personalized care strategies. For instance, in autism, Sensory Integration Therapy (SIT) has shown promise in improving sensory regulation and responses to stimuli (Schaaf et al., 2014). Similar approaches could be adapted to address the sensory profiles seen in the NDP of BD, such as sensory modulation or occupational therapy. Finally, NDP-informed screening could contribute to earlier identification of at-risk individuals and to preventive or remediation strategies. Atypical sensory processing in ASD has been associated with poorer adaptive functioning, independently of cognitive ability (Neufeld et al., 2021, underscoring the role of sensory processing in everyday functioning. Given the chronic functional impairments observed in BD, adding a sensory based approach to their treatment could be very useful, and the NDP seems to be a precise tool for detection.

Overall, the present findings contribute to the comprehension of sensory processing alterations in BD, but have some limitations. Although acceptable, the sample size was moderate and some measures had missing data which may have limited the statistical power in our analyses. In other words, we found significant relationships between NDP and SP, but the causal relationship from NDP to SP requires more data (supplementary Table S1). Also, the cross-sectional nature of this study limits causal inference; future longitudinal research is needed to clarify the developmental trajectory of sensory alterations in BD. Sensory processing was assessed using a self-report questionnaire, which, although validated, may be influenced by self-perception biases or residual mood symptoms, even in euthymic individuals. Using another SP measure could strengthen the reliability of the sensory profiles found. Additionally, the potential effects of pharmacological treatment such as antidepressants, mood stabilizers, or benzodiazepines were not systematically examined.

Taken together, these findings contribute to the understanding of sensory system functioning in the NDP of BD. Despite the growing recognition of SP in brain functioning, research in this area remains limited. Existing studies often differ in terms of diagnostic subtypes (e.g., BD type I and II, often mixed samples) and in mood state during assessment, adding to the complexity posed by the intrinsic heterogeneity of BD. Unlike prior studies conducted during acute episodes (Parker et al., 2017; Shaffer et al., 2018), our study assessed SP during euthymia. This approach allowed minimizing state-dependent effects while highlighting trait-like sensory alterations associated with the NDP.

In conclusion, this study provides novel evidence that increased sensory processing alterations, particularly increased *low registration, sensory sensitivity,* and *sensation avoiding* behaviors, are significantly associated with the NDP phenotype in euthymic individuals with BD. These findings support the hypothesis that sensory processing dimensions may function as stable, trait-like markers of neurodevelopmental vulnerability in BD. Furthermore, our study validated the NDP as a relevant stratification approach in BD in regard to sensory processing. These findings support the incorporation of developmental and sensory phenotyping into BD research and clinical practice to refine prognostic models and to identify candidates for sensory-focused interventions. By highlighting the relevance of these sensory patterns beyond acute mood episodes, our results offer promising avenues for earlier identification, more personalized clinical interventions, and ultimately, improved long-term functional outcomes in this population. More studies are needed to validate our findings and to explore their implications for long-term functional outcomes in BD.

## Data Availability

De-identified participant data and analysis code are
available from the corresponding author on reasonable request.

## Declarations

### Ethics approval and consent to participate

Approved by Ethics Committee of Aix-Marseille University (ref. no. 2025-03-13-09). All participants provided written informed consent.

### Consent for publication

All authors consent to publication.

### Availability of data and materials

De-identified participant data and analysis code are available from the corresponding author on reasonable request.

## Acknowledgements

We thank the FACE-BD team and participants for their contribution.

## Supplemental material

**Supplementary Figure S1.**
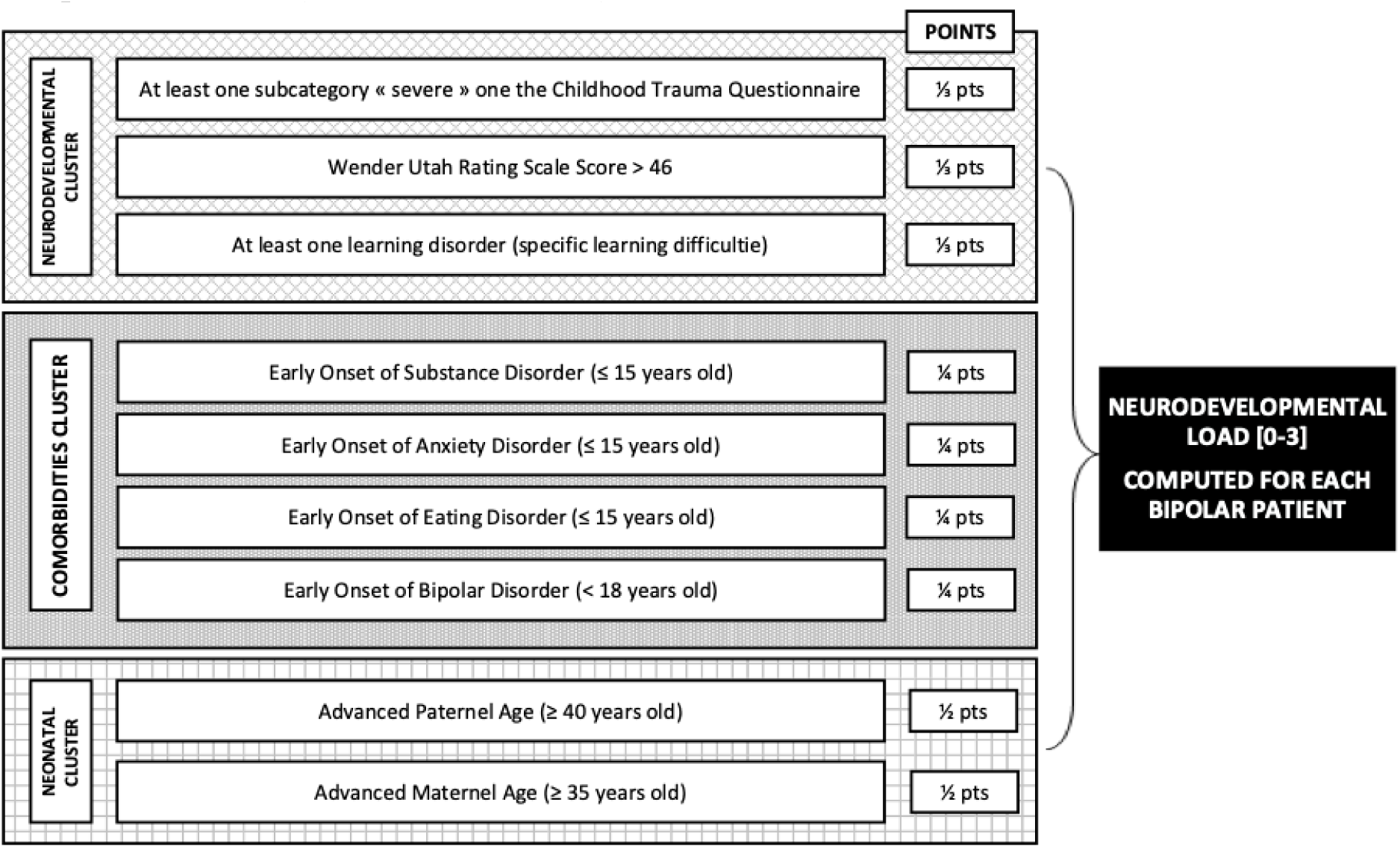
The three clusters establishing the neurodevelopmental load score in bipolar disorders (Lefrère, et al., 2024).

**Supplementary Figure S2.**
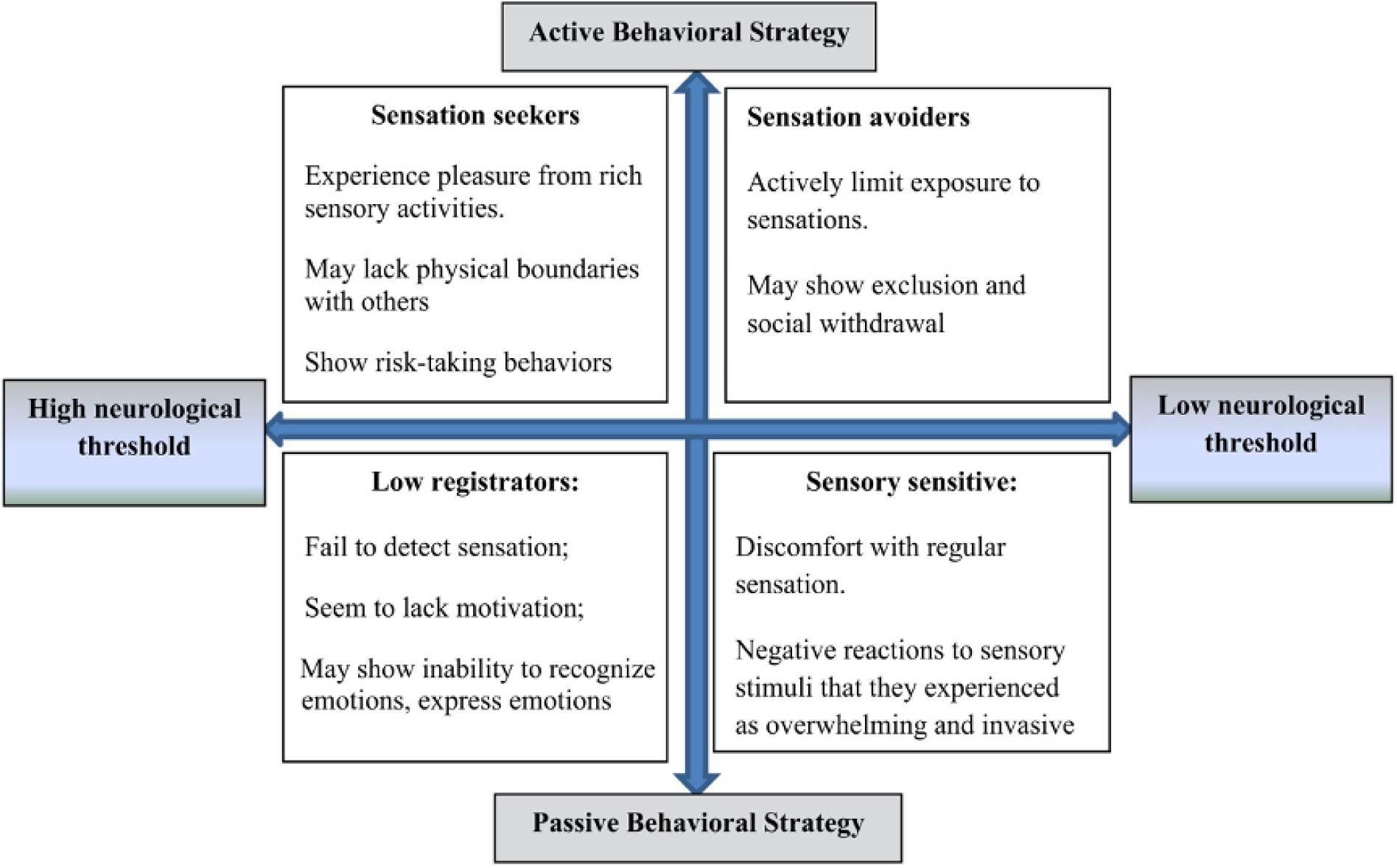
Dunn’s sensory processing model (1997).

**Supplementary Figure S3.**
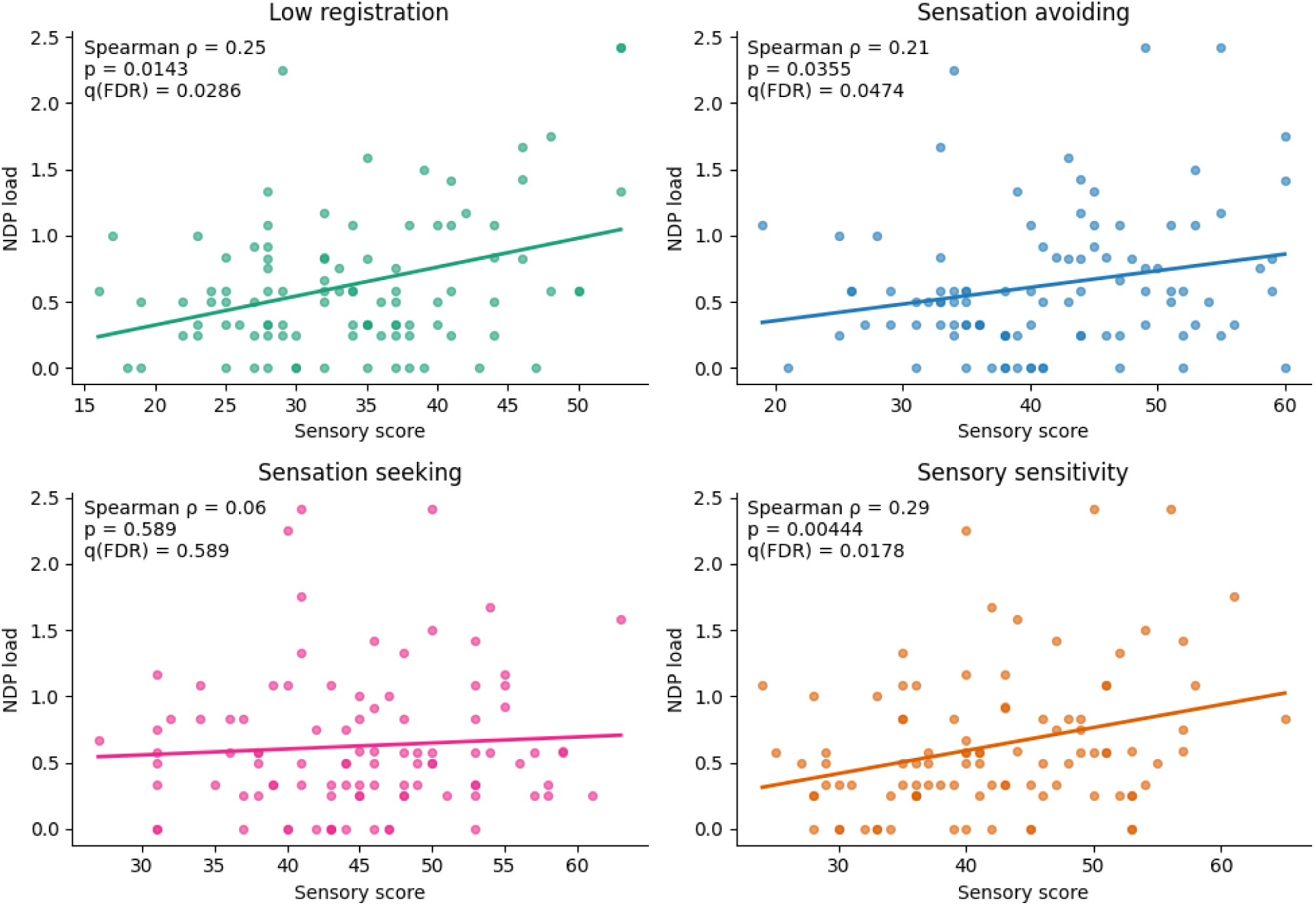
Associations between sensory processing profiles and neurodevelopmental polarity load in bipolar disorder Scatterplots depict the relationships between Adolescent/Adult Sensory Profile quadrant scores (Low registration, Sensation avoiding, Sensation seeking, Sensory sensitivity) and the NDP load in bipolar participants (BD and BD-ND). Each panel represents one sensory quadrant. Solid lines indicate least-squares regression fits for visualization purposes. Spearman rank correlations (ρ) are reported with corresponding raw p-values and false discovery rate (FDR)–corrected q-values (Benjamini–Hochberg). Significant associations after FDR correction were observed for Low registration, Sensation avoiding, and Sensory sensitivity, whereas Sensation seeking showed no significant association with NDP load.

**Supplementary Table S1.**
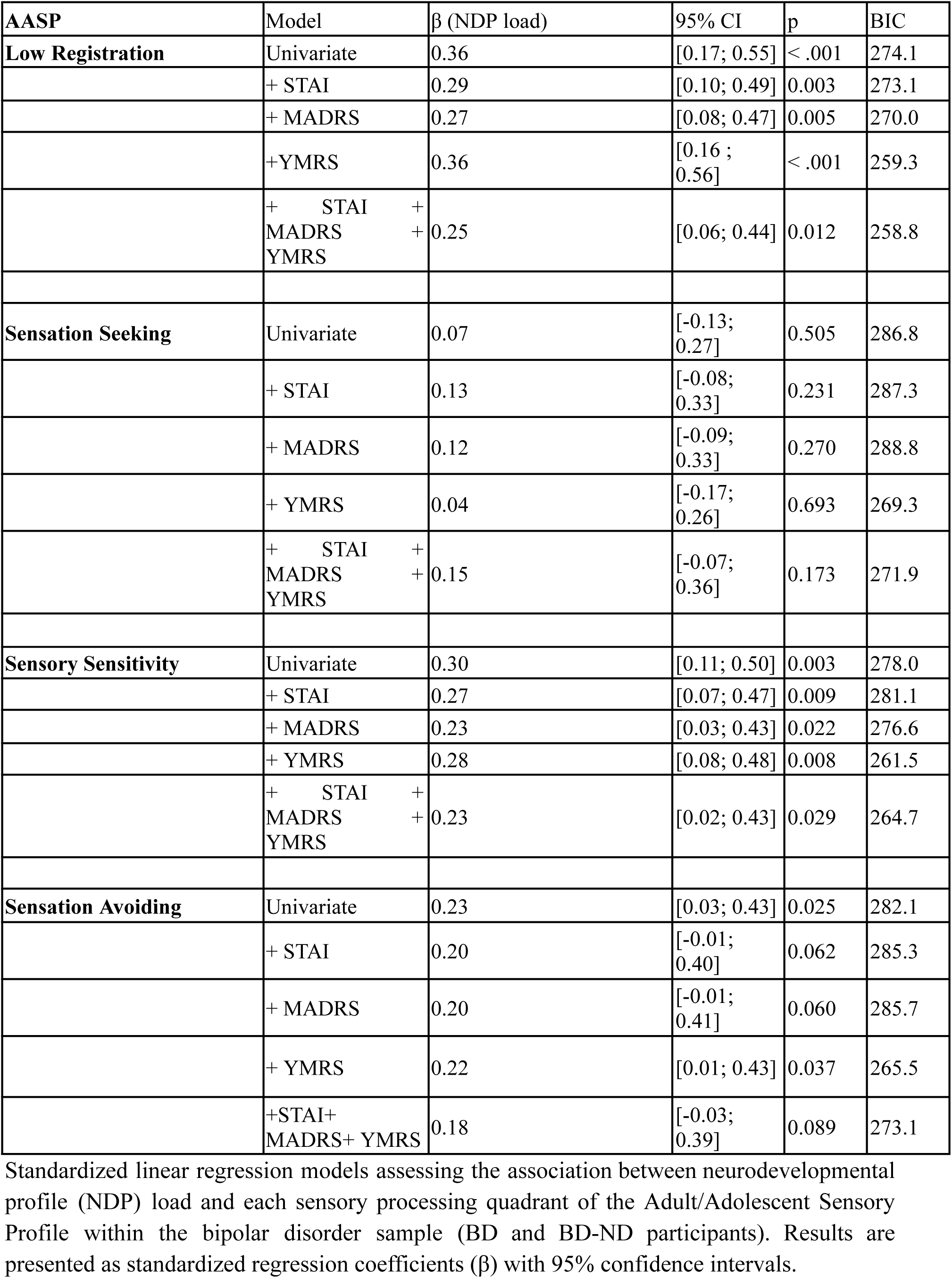
Association between NDP load and sensory processing quadrants in the bipolar disorder sample.

| AASP | Model | $\beta$ (NDP load) | 95% CI | p | BIC |
| --- | --- | --- | --- | --- | --- |
| **Low Registration** | Univariate | 0.36 | [0.17; 0.55] | < .001 | 274.1 |
|  | + STAI | 0.29 | [0.10; 0.49] | 0.003 | 273.1 |
|  | + MADRS | 0.27 | [0.08; 0.47] | 0.005 | 270.0 |
|  | +YMRS | 0.36 | [0.16 ; 0.56] | < .001 | 259.3 |
|  | + STAI +<br>MADRS +<br>YMRS | 0.25 | [0.06; 0.44] | 0.012 | 258.8 |
| **Sensation Seeking** | Univariate | 0.07 | [-0.13; 0.27] | 0.505 | 286.8 |
|  | + STAI | 0.13 | [-0.08; 0.33] | 0.231 | 287.3 |
|  | + MADRS | 0.12 | [-0.09; 0.33] | 0.270 | 288.8 |
|  | + YMRS | 0.04 | [-0.17; 0.26] | 0.693 | 269.3 |
|  | + STAI +<br>MADRS +<br>YMRS | 0.15 | [-0.07; 0.36] | 0.173 | 271.9 |
| **Sensory Sensitivity** | Univariate | 0.30 | [0.11; 0.50] | 0.003 | 278.0 |
|  | + STAI | 0.27 | [0.07; 0.47] | 0.009 | 281.1 |
|  | + MADRS | 0.23 | [0.03; 0.43] | 0.022 | 276.6 |
|  | + YMRS | 0.28 | [0.08; 0.48] | 0.008 | 261.5 |
|  | + STAI +<br>MADRS +<br>YMRS | 0.23 | [0.02; 0.43] | 0.029 | 264.7 |
| **Sensation Avoiding** | Univariate | 0.23 | [0.03; 0.43] | 0.025 | 282.1 |
|  | + STAI | 0.20 | [-0.01; 0.40] | 0.062 | 285.3 |
|  | + MADRS | 0.20 | [-0.01; 0.41] | 0.060 | 285.7 |
|  | + YMRS | 0.22 | [0.01; 0.43] | 0.037 | 265.5 |
|  | +STAI+<br>MADRS+ YMRS | 0.18 | [-0.03; 0.39] | 0.089 | 273.1 |

Standardized linear regression models assessing the association between neurodevelopmental profile (NDP) load and each sensory processing quadrant of the Adult/Adolescent Sensory Profile within the bipolar disorder sample (BD and BD-ND participants). Results are presented as standardized regression coefficients ( $\beta$ ) with 95% confidence intervals.

**Supplementary Table S2.**
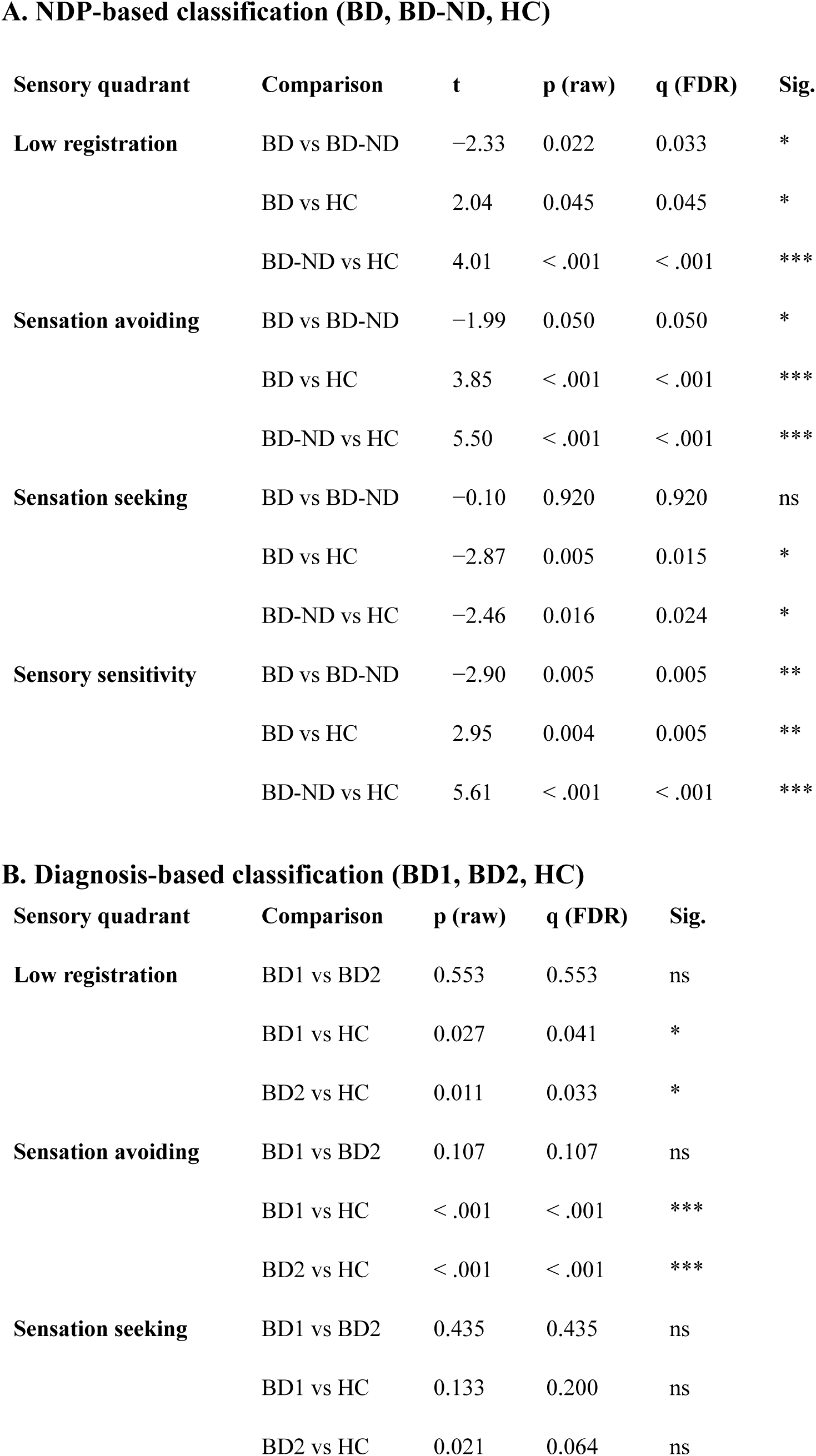
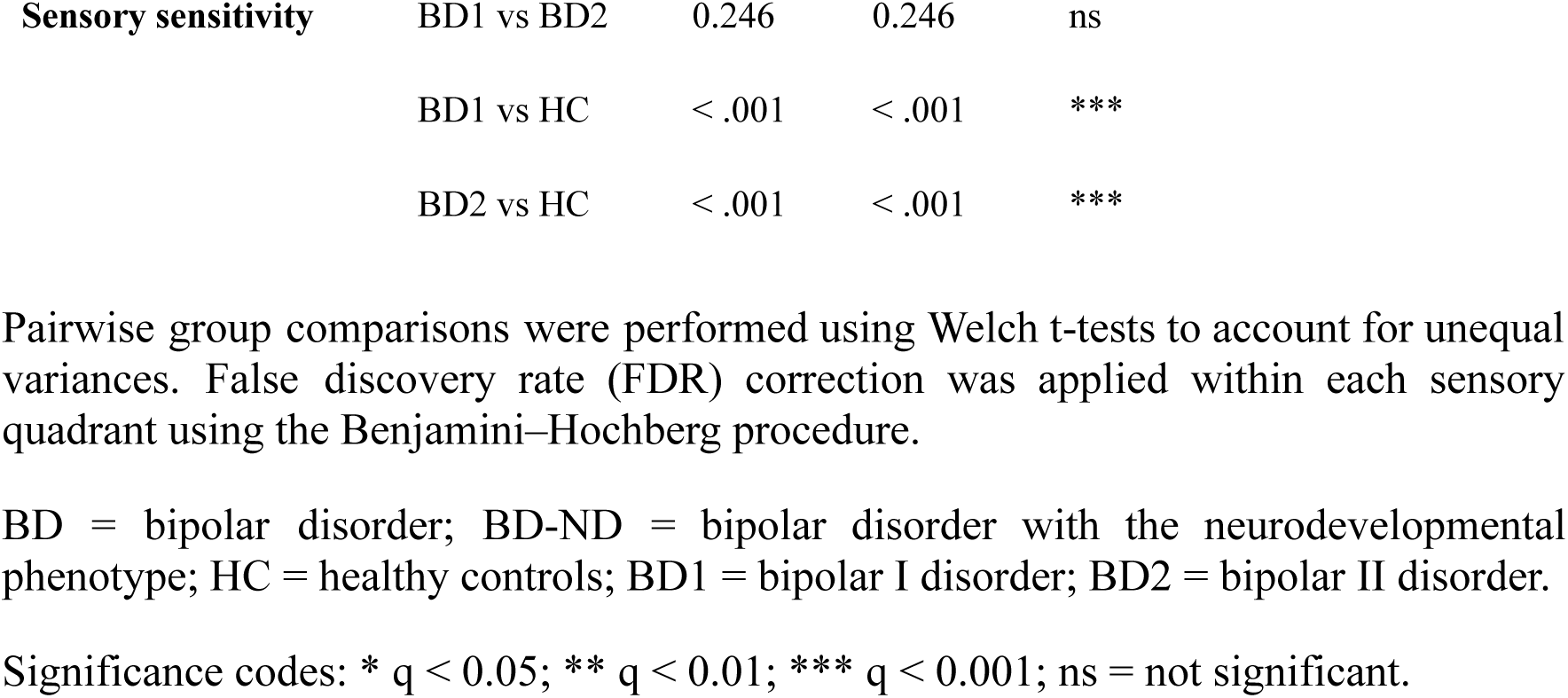
Post-hoc pairwise comparisons of sensory processing quadrants across diagnostic and NDP-based classifications.

